# Association of Age-Related Macular Degeneration With Mortality: A Propensity Score-Matched Analysis

**DOI:** 10.1101/2022.01.18.22269511

**Authors:** Yifan Chen, Yueye Wang, Xianwen Shang, Wei Wang, Zhuoting Zhu

**Affiliations:** Guangdong Eye Institute, Department of Ophthalmology, Guangdong Provincial People’s Hospital, Guangdong Academy of Medical Sciences, Guangzhou, 510080, China; John Radcliffe Hospital, Oxford University Hospitals NHS Foundation Trust, Oxford, UK; State Key Laboratory of Ophthalmology, Zhongshan Ophthalmic Center, Sun Yat-sen University, Guangzhou, China

**Keywords:** age-related macular degeneration, all-cause mortality, cause-specific mortality

## Abstract

**Purpose:** To investigate the association between age-related macular degeneration (AMD) and 10-year all-cause and cause-specific mortality using a large-scale population-based sample.

**Methods:** Data from the 2005-2008 cycles of the National Health and Nutrition Examination Survey were used to assess the risk of mortality in relation to AMD in a propensity score-matched cohort. AMD status was assessed by retinal images with the standardized grading scheme. Mortality data until 31st December 2015 were derived from mortality archives. Cox proportional hazards regression models were used to estimate hazard ratios (HRs) and 95% confidence intervals (CIs) for survival.

**Results:** A total of 4691 participants were included. After a median follow-up of 8.42 (IQR: 7.58-9.67) years, 698 participants died. Participants with any AMD had an increased risk of all-cause mortality (HR, 2.02; 95% CI, 1.37-2.98). Similar results were observed for early (HR, 1.93; 95% CI, 1.31-2.85) and late AMD (HR, 4.29; 95% CI, 2.10-8.79). For cause-specific mortality, any (HR, 2.17; 95% CI, 1.39-3.39), early (HR, 2.18; 95% CI, 1.36-3.51), and late AMD (HR, 3.95; 95% CI, 1.65-9.46) were associated with significantly higher mortality due to causes other than cardiovascular disease (CVD) or cancer. Late AMD independently predicted a higher risk of CVD mortality (HR, 2.48; 95% CI, 1.32-4.65).

**Conclusions:** The current study showed that any, early, and late AMD were associated with increased risks of all-cause mortality and mortality due to causes other than CVD or cancer. In addition, we found that late AMD was associated with increased risks of CVD mortality.

**Synopsis:** Late macular degeneration independently predicted higher cardiovascular disease mortality. Any, early and late age-related macular degeneration were associated with higher all-cause mortality and mortality due to causes other than cardiovascular disease or cancer.

## Introduction

Age-related macular degeneration (AMD) is one of the leading causes of visual impairment and blindness worldwide.^1^ The number of people with AMD globally is projected to increase to 288 million in 2040.^2^ In the United States, the number of people with AMD is estimated to reach 2.95 million in 2020.^3^ As the population ages, the prevalence of AMD is likely to increase further, leading to significant economic and societal costs.^4^

Accumulating evidence has suggested that AMD is an independent risk factor for all-cause mortality.^5–10^ However, the results for its association with cause-specific mortality remain inconsistent.^11–20^ Some studies have reported that AMD was associated with an increased risk of CVD mortality,^5,6,8^ while others did not find any significant association between AMD and CVD mortality.^11,12,19^ Investigators have previously hypothesized that the similar risk profile shared by AMD and CVD^21–23^, as well as the potential thromboembolic events secondary to intravitreal anti-vascular endothelial growth factor (anti-VEGF) treatment^24,25^ may increase the risk of CVD mortality among patients with AMD.

Our previous study published in 2018 found that only late AMD increased the risk of 5-year all-cause and non-CVD non-cancer mortality, while no significant association was found between AMD and CVD mortality using data from the National Health and Nutrition Examination Survey (NHANES).^26^ The availability of 10-year mortality data empowers us with longer follow-up and more statistical power to investigate the long-term mortality among patients with AMD.

The propensity score reflects the probability of being exposed based on baseline covariates^27^. As a popular method used for controlling confounders, propensity score matching works with prevalence in giving unbiased estimate in observational studies with multiple covariates and rare outcome^28,29^. Thus, in this study, propensity score matching was employed instead of conventional parsimonious regression models by matching participants with similar distribution of confounders, to maximum eliminate potential bias. Therefore, we aim to investigate the association of AMD with 10-year all-cause and cause-specific mortality using a propensity score-matched method.

## Methods

### Sample and Population

The NHANES is conducted by the National Center for Health Statistics (NCHS) of the Centers for Disease Control and Prevention. It uses stratified multistage sampling methods, described in detail elsewhere.^30^ Briefly, all participants completed health-related interviews and examinations in each cycle of the NHANES, which was conducted every two years. In this survey analysis, we analyzed data from the 2005-2008 cycles of the NHANES. NHANES protocols were reviewed and approved by the NCHS research ethics review committee and all participants provided written informed consent.

### AMD Identification

During the 2005-2008 cycles of NHANES, retinal images were collected from participants aged 40 years and older using an ophthalmic digital imaging system (CR6-45NM; Canon USA, Inc) and digital camera (EOS 10D; Canon USA, Inc). Fundus images were graded at the University of Wisconsin, Madison, according to the modified Wisconsin Age-Related Maculopathy Grading Classification Scheme.^31,32^ Early AMD was defined as signs of drusen with a grid area of more than a 500 µm circle and/or pigmentary abnormalities. Late AMD was defined as the presence of exudative or geographic atrophy signs. The AMD status was decided based on the more severely affected eye.

### Mortality Data

Mortality data were derived from the 2020 public-access linked mortality archives.^33^ The International Statistical Classification of Diseases and Related Health Problems, Tenth Revision (ICD-10) codes were used to determine causes of deaths. Codes I00 to I09, I11, I13, and I20 to I51 (diseases of heart) and I60 to I69 (cerebrovascular disease) were defined as death attributable to CVD. ICD-10 codes C00 to C97 were defined as cancer mortality. Any other causes of death were considered death due to causes other than CVD or cancer. Participants were considered alive if not matched with death certificates. Time to death was counted from baseline to date of death or the 31^st^ of December, 2015, whichever came first.

### Confounding Variables

Information on demographics, health-related behaviors, and other characteristics was collected by in-person interviews and examinations. The confounding variables considered in this analysis include age, gender, ethnicity, education, marital status, family income, smoking status, alcohol consumption, diabetes mellitus, hypertension, high cholesterol, body mass index, C-reactive protein level, walking disability, self-rated health status, depressive symptoms, coexisting ocular diseases, history of cardiovascular diseases and history of cancer. Details on how these confounding variables were determined were described in a previous paper.^26,34–37^ The exclusion on confounding factors was done if any one of the factors were missing.

### Statistical Analysis

Data from the 2005-2006 and 2007-2008 cycles of NHANES were combined and analysed accounting for the complex and stratified designed based on the NHANES analytic and reporting guidelines.^38^ In the present analysis, we assessed the risk of mortality in relation to AMD in a propensity score-matched cohort. Logistic regression models were used to create the propensity score and the balance of covariates in models was tested. Inverse probability of treatment weighting method based on the synthesized propensity score was used to match participants with and without AMD. Unpaired t-test and design-adjusted Rao-Scott Pearson χ^2^test were used for the comparison of continuous and categorical variables, respectively. Hazard ratios (HRs) and their 95% confidence interval (95% CI) were estimated by Cox proportional hazards regression models. Sensitivity analyses that excluding the participants with cancer at baseline were further performed to validate the robustness of our findings. All analyses were carried out in Stata (Version 14; Stata Corp, College Station, TX). A P value of less than 0.05 was considered to be significant.

## Results

### Study Population

After excluding 1193 participants who had missing information on the AMD status and/or survival status, and 913 participants who had missing information on confounding factors, 4691 participants were identified. Excluded participants were more likely to be elderly, of female gender, non-Hispanic black and other ethnicities, lower educational level, unmarried status, below poverty line, and lifetime abstainer/former drinker. They also tended to have comorbidities history, coexisting ocular diseases, walking disability, and poorer general health status (Supplement Table 1).

There were 360 participants with AMD and 4331 participants without AMD. There was no significant difference in baseline characteristics between AMD and non-AMD participants after propensity score matching (Table 1).

**Table 1.** Characteristics of Participants With And Without AMD in Propensity Score-Matched Cohorts.

| Characteristics | No. (%) |  | P Value<sup>a</sup> |
| --- | --- | --- | --- |
|  | Non-AMD | AMD |  |
| Age, yrs |  |  |  |
| 40-49 | 1263 (34.9) | 24 (43.5) | 0.312 |
| 50-59 | 1100 (31.0) | 39 (24.2) |  |
| 60-69 | 1084 (18.6) | 80 (17.8) |  |
| 70-79 | 644 (11.2) | 107 (9.64) |  |
| ≥80 | 240 (4.33) | 110 (4.94) |  |
| Gender |  |  |  |
| Male | 2185 (48.0) | 187 (52.5) | 0.408 |
| Female | 2146 (52.0) | 173 (47.5) |  |
| Race |  |  |  |
| Non-Hispanic white | 2347 (78.4) | 258 (84.7) | 0.050 |
| Non-Hispanic black | 871 (8.96) | 31 (4.29) |  |
| Mexican American | 689 (5.32) | 42 (5.90) |  |
| Other | 424 (7.32) | 29 (5.16) |  |
| Education |  |  |  |
| Less than high school | 1210 (17.1) | 100 (17.1) | 0.996 |
| High school and over | 3121 (82.9) | 260 (82.9) |  |
| Marital status |  |  |  |
| Unmarried and other | 1481 (30.2) | 169 (23.6) | 0.142 |
| Married/with a partner | 2850 (69.8) | 191 (76.4) |  |
| Poverty income ratio (PIR) |  |  |  |
| Below poverty (<1) | 640 (8.46) | 55 (7.57) | 0.723 |
| At or above poverty ($\geq 1$) | 3691 (91.5) | 305 (92.4) | |
| Smoking status |  |  |  |
| Never | 2057 (48.7) | 164 (41.5) | 0.292 |
| Former | 1376 (30.9) | 145 (39.7) |  |
| Current | 898 (20.4) | 51 (18.8) |  |
| Alcohol consumption |  |  |  |
| Lifetime abstainer/former drinker | 1025 (19.8) | 100 (17.4) | 0.588 |
| Current drinker | 2362 (55.5) | 185 (60.5) |  |
| ($\leq 3$ drinks/w) | | | |
| Current drinker | 944 (24.7) | 75 (22.1) |  |
| (> 3 drinks/w) |  |  |  |
| Diabetes mellitus |  |  |  |
| No | 3535 (87.0) | 299 (89.7) | 0.296 |
| Yes | 796 (13.0) | 61 (10.3) |  |
| Hypertension |  |  |  |
| No | 2243 (56.7) | 123 (62.1) | 0.440 |
| Yes | 2088 (43.3) | 237 (37.9) |  |
| High cholesterol |  |  |  |
| No | 2653 (62.8) | 214 (58.4) | 0.468 |
| Yes | 1678 (37.2) | 146 (41.6) |  |
| BMI |  |  |  |
| <18.5 kg/m<sup>2</sup> | 62 (1.30) | 3 (0.21) | 0.103 |
| 18.5-30.0 kg/m<sup>2</sup> | 2594 (61.8) | 249 (68.6) |  |
| $\geq 30.0$ kg/m<sup>2</sup> | 1675 (36.9) | 108 (31.2) | |
| High C-reactive protein |  |  |  |
| No | 3839 (89.6) | 315 (92.6) | 0.317 |
| Yes | 492 (10.4) | 45 (7.40) |  |
| Depressive symptom |  |  |  |
| No | 3967 (93.0) | 337 (90.6) | 0.509 |
| Yes | 364 (7.05) | 23 (9.40) |  |
| Coexisting ocular diseases |  |  |  |
| No | 3334 (81.5) | 204 (84.0) | 0.450 |
| Yes | 997 (18.5) | 156 (16.0) |  |
| Walking disability |  |  |  |
| No | 3950 (92.8) | 300 (91.6) | 0.532 |
| Yes | 381 (7.20) | 60 (8.44) |  |
| Self-rated health |  |  |  |
| Poor/Fair | 1054 (17.6) | 92 (17.1) | 0.896 |
| Good/Excellent | 3277 (82.4) | 268 (82.9) |  |
| History of congestive heart failure |  |  |  |
| No | 4156 (97.0) | 339 (94.2) | 0.314 |
| Yes | 175 (3.02) | 21 (5.76) |  |
| History of heart attack |  |  |  |
| No | 4094 (95.4) | 322 (92.9) | 0.424 |
| Yes | 237 (4.58) | 38 (7.10) |  |
| History of stroke |  |  |  |
| No | 4157 (96.3) | 315 (96.5) | 0.802 |
| Yes | 174 (3.71) | 45 (3.46) |  |
| History of cancer |  |  |  |
| No | 3818 (88.2) | 294 (91.4) | 0.198 |
| Yes | 513 (11.8) | 66 (8.56) |  |

Abbreviations: AMD, age-related macular degeneration; BMI, body mass index.
<sup>a</sup> All P values were calculated using the design-adjusted Rao-Scott Pearson $\chi^2$ test for categorical variables.

**Table 2.** Risk for All-Cause Mortality and Specific-Cause Mortality in Participants With AMD Compared With Non-AMD After Propensity Score Matching

|  | All-Cause Mortality |  | CVD Mortality |  | Cancer Mortality |  | Non-CVD/Non-cancer Mortality |  |
| --- | --- | --- | --- | --- | --- | --- | --- | --- |
|  | Events/Number | HR (95% CI)<sup>a</sup> | Events/Number | HR (95% CI)<sup>a</sup> | Events/Number | HR (95% CI)<sup>a</sup> | Events/Number | HR (95% CI)<sup>a</sup> |
| AMD status |  |  |  |  |  |  |  |  |
| No AMD | 571/4331 | Reference | 110/4331 | Reference | 142/4331 | Reference | 319/4331 | Reference |
| Any AMD | 127/360 | **2.02 (1.37-2.98)** | 19/360 | 1.42 (0.69-2.92) | 22/360 | 1.42 (0.72-2.82) | 86/360 | **2.17 (1.39-3.39)** |
| Early AMD | 104/318 | **1.93 (1.31-2.85)** | 15/318 | 1.27 (0.58-2.82) | 20/318 | 1.45 (0.73-2.89) | 69/318 | **2.18 (1.36-3.51)** |
| Late AMD | 23/42 | **4.29 (2.10-8.79)** | 4/42 | **2.48 (1.32-4.65)** | 2/42 | 0.44 (0.06-3.27)<sup>b</sup> | 17/42 | **3.95 (1.65-9.46)** |

Abbreviations: AMD, age-related macular degeneration; CVD, cardiovascular disease; CI, confidence interval; HR, hazard ratio.
<sup>a</sup> Adjusted for age, gender, race, education, marital status, family income, smoking status, alcohol consumption, diabetes mellitus, hypertension, high cholesterol, BMI, high CRP, depressive symptom, coexisting ocular diseases, walking disability, self-rated health, history of cardiovascular diseases, and cancer.
<sup>b</sup> Adjusted for age and gender.
Boldface indicates statistical significance.

### All-cause mortality

After a median duration of 8.42 (IQR: 7.58-9.67) years, a total of 698 participants died from all causes. After adjustments for confounding factors, the Cox proportional hazards regression model indicated that any AMD was associated with increased all-cause mortality (HR, 2.02; 95% CI, 1.37-2.98). Early AMD (HR, 1.93; 95% CI, 1.31-2.85) and late AMD (HR, 4.29; 95% CI, 2.10-8.79) were also associated with significantly higher all-cause mortality compared to the group without AMD.

### Cause-specific mortality

Of all deaths, 129, 164, and 405 were attributable to CVD, cancer, and non-CVD non-cancer causes, respectively. After multiple adjustments, Cox proportional hazards regression model showed that any AMD (HR, 2.17; 95% CI, 1.39-3.39), early AMD (HR, 2.18; 95% CI, 1.36-3.51) and late AMD (HR, 3.95; 95% CI, 1.65-9.46) were significantly associated with higher risks of mortality due to causes other than CVD or cancer. In the fully adjusted model, we found that late AMD independently predicted a higher risk of CVD mortality (HR, 2.48; 95% CI, 1.32-4.65).

### Sensitivity analysis

Sensitivity analyses that excluding the participants with cancer at baseline were further performed to validate the robustness of our findings. Similar results were noted (Supplement Table 2).

## Discussion

The present study showed that any, early and late AMD were significantly associated with increased risks of all-cause mortality and mortality due to causes other than CVD or cancer, which was consistent with our previous study^26^ with a 5-year follow-up duration. In addition, we found that late AMD is associated with increased CVD mortality.

Consistent evidence on the association between late AMD and increased risks of all-cause mortality has been reported.^6,12,26,39,40^ Whereas conflicting results were found on the associations of early or any AMD with all-cause mortality.^6–8,11–13,15,17,20,39^ These conflicting results might be due to the heterogeneity in the study population, detection of AMD, statistical methods, and confounding factors adjusted for. The present analysis indicated that any form of AMD was associated with increased risks of mortality, suggesting the presence of AMD might be a biomarker of ageing and frailty. Even though the exact mechanism for the significant association between AMD and mortality remains unclear, growing evidence has indicated some common pathways shared between the ageing process and the pathogenesis of AMD, such as increased oxidative stress and chronic inflammation.^41–43^

The present study found that participants with late AMD had significantly higher CVD mortality. Growing evidence has demonstrated a close relationship between AMD and increased CVD mortality.^5,6,8,11,12,34,44^ Two recent systematic reviews found that late AMD was significantly associated with increased CVD mortality with an estimated HR of 1.46 and 1.28 respectively.^39,40^ The association between late AMD and CVD mortality might be explained by their common pathogenesis pathways (e.g., local inflammation and oxidative stress) and risk factors (e.g., age and smoking).^35,45,46^ The ‘hallmark’ of AMD, drusen, closely resembled atherosclerotic plaques in composition.^46^ In addition, the use of anti-VEGF to treat neovascular AMD might also increase CVD risks leading to higher CVD mortality.^24,25,47,48^

Consistent with previous studies,^10,11^ we also found a strong association between AMD and non-CVD non-cancer mortality. Several hypotheses may explain this. Firstly, participants with AMD may be more generally frail. Secondly, increasing evidence suggested that AMD was significantly associated with neurodegenerative diseases (e.g., Alzheimer’s disease).^49–51^ Last but not least, participants with AMD might be more likely to suffer from functional disabilities and social deprivation due to impaired vision.^52–54^

The present study has the advantage of a large-scale nationally sample, long-term follow-up and the use of propensity score matching method to eliminate confounding effects, which could fairly imitate the settings of random clinical trials. Moreover, the high consistency among results of the multiple sensitivity analyses and the primary analysis strongly supported the validity of our findings. Several limitations should be considered. Firstly, the relatively small number of participants with late AMD preventing us from conducting further subgroup analyses. Large-scale studies are needed to corroborate our findings. Secondly, the AMD status being assessed at a single time point, the consecutive trend could only be inferred. Thirdly, though we have included and matched most of the previously established confounders, less precise estimates from propensity scores may be drawn due to potential unmatched baseline characteristics. Last but not the least, the differences in baseline characteristics between excluded participants and included participants might bias the AMD-mortality associations.

## Conclusion

In summary, the present study showed that any, early, and late AMD were associated with increased risks of all-cause mortality and mortality due to causes other than CVD or cancer. Furthermore, we also found that late AMD independently predicted a higher risk of CVD mortality. Our results highlight the importance of early detection and management of AMD as this helps to identify the vulnerable group of patients who have increased long-term mortality.

## Data Availability

All data produced in the present study are available upon reasonable request to the authors

https://wwwn.cdc.gov/nchs/nhanes/default.aspx

## 3) Other Acknowledgement

None.

## Author Contributions

Study concept and design: Yifan Chen, Wei Wang, Zhuoting Zhu;

Acquisition, analysis, or interpretation: Xianwen Shang, Wei Wang, Zhuoting Zhu;

Drafting of the manuscript: Yifan Chen, Yueye Wang;

Critical revision of the manuscript for important intellectual content: Xianwen Shang,

Wei Wang, Zhuoting Zhu;

Statistical analysis: Xianwen Shang, Wei Wang, Zhuoting Zhu;

Obtained funding: Wei Wang, Zhuoting Zhu;

Administrative, technical, or material support: Wei Wang, Zhuoting Zhu;

Study supervision: Xianwen Shang, Wei Wang, Zhuoting Zhu;

## Abbreviations and Acronyms

AMD: age-related macular degeneration
anti-VEGF: anti-vascular endothelial growth factor
NHANES: National Health and Nutrition Examination Survey
NCHS: National Center for Health Statistics
ICD-10: International Statistical Classification of Diseases and Related Health Problems, Tenth Revision
HR: Hazard ratios
CI: confidence interval

