## Supplementary Table 1-2 for "Association of Age-Related Macular Degeneration With Mortality: A Propensity Score-Matched Analysis"

Supplement Table 1. Characteristics of Participants Included and Excluded in the Current Analysis.

| Characteristics | No. (%) | | P Value[a] |
|---|---|---|---|
| | Excluded | Included | |
| Age, yrs | | | |
| 40-49 | 455(29.3) | 1287(35.1) | **<0.001** |
| 50-59 | 383(23.7) | 1139(30.7) | |
| 60-69 | 460(17.6) | 1164(18.6) | |
| 70-79 | 411(15.1) | 751(11.0) | |
| ≥80 | 297(14.3) | 350(4.61) | |
| Gender | | | |
| Male | 1001(44.5) | 2372(48.0) | **0.028** |
| Female | 1105(55.5) | 2319(52.0) | |
| Race | | | |
| Non-Hispanic white | 941(66.9) | 2605(78.6) | **<0.001** |
| Non-Hispanic black | 567(15.6) | 902(8.76) | |
| Mexican American | 317(5.90) | 731(5.39) | |
| Other | 281(11.6) | 453(7.29) | |
| Education | | | |
| Less than high school | 831(26.2) | 1310(17.2) | **<0.001** |
| High school and over | 1275(73.8) | 3381(82.8) | |
| Marital status | | | |
| Unmarried and other | 926(38.1) | 1650(30.0) | **<0.001** |
| Married/with a partner | 1174(61.9) | 3041(70.0) | |
| Poverty income ratio (PIR) | | | |
| Below poverty (<1) | 349(15.2) | 695(8.47) | **<0.001** |
| At or above poverty (≥1) | 1228(84.8) | 3996(68.1) | |
| Smoking status | | | |
| Never | 1074(51.3) | 2221(48.6) | 0.163 |
| Former | 636(28.7) | 1521(31.1) | |

| | | | |
|---|---|---|---|
| Current | 389(20.0) | 949(20.4) | |
| Alcohol consumption | | | |
| Lifetime abstainer/former drinker | 510(29.3) | 1125(19.8) | **<0.001** |
| Current drinker (≤ 3 drinks/w) | 786(51.9) | 2547(55.6) | |
| Current drinker (> 3 drinks/w) | 255(18.8) | 1019(24.6) | |
| Diabetes mellitus | | | |
| No | 1291(77.7) | 3834(86.9) | **<0.001** |
| Yes | 539(22.3) | 857(13.1) | |
| Hypertension | | | |
| No | 767(49.1) | 2366(56.8) | **<0.001** |
| Yes | 1014(50.9) | 2325(43.2) | |
| High cholesterol | | | |
| No | 1075(59.0) | 2867(62.7) | 0.053 |
| Yes | 738(41.0) | 1824(37.3) | |
| BMI | | | |
| <18.5 kg/m$^2$ | 38(2.21) | 65(1.20) | 0.102 |
| 18.5-30.0 kg/m$^2$ | 1187(62.3) | 2843(62.0) | |
| ≥ 30.0 kg/m$^2$ | 722(35.5) | 1783(36.8) | |
| High C-reactive protein | | | |
| No | 1518(88.3) | 4154(89.6) | 0.205 |
| Yes | 220(11.7) | 537(10.5) | |
| Depressive symptom | | | |
| No | 1362(92.1) | 4304(92.9) | 0.485 |
| Yes | 137(7.87) | 387(7.09) | |
| Coexisting ocular diseases | | | |
| No | 581(58.9) | 3538(81.3) | **<0.001** |
| Yes | 527(41.1) | 1153(18.7) | |

| | | | |
|---|---|---|---|
| Walking disability | | | |
| No | 1645(81.9) | 4250(92.7) | **<0.001** |
| Yes | 461(18.1) | 441(7.30) | |
| Self-rated health | | | |
| Poor/Fair | 584(28.9) | 1146(17.6) | **<0.001** |
| Good/Excellent | 994(71.1) | 3545(82.4) | |
| History of congestive heart failure | | | |
| No | 1941(94.1) | 4495(96.9) | **<0.001** |
| Yes | 165(5.92) | 196(3.08) | |
| History of heart attack | | | |
| No | 1922(92.7) | 4416(95.4) | **<0.001** |
| Yes | 184(7.29) | 275(4.58) | |
| History of stroke | | | |
| No | 1915(92.7) | 4472(96.3) | **<0.001** |
| Yes | 191(7.28) | 219(3.73) | |
| History of cancer | | | |
| No | 1804(84.9) | 4112(88.2) | **0.007** |
| Yes | 302(15.1) | 579(11.8) | |

Abbreviations: AMD, age-related macular degeneration; BMI, body mass index.

[a] All P values were calculated using the design-adjusted Rao-Scott Pearson $\chi 2$ test for categorical variables.

Supplement Table 2. Sensitivity Analysis on the Risk for All-Cause Mortality and Specific-Cause Mortality in Participants With AMD Compared With Non-AMD After Propensity Score Matching

| | **HR (95% CI)[a]** | | | |
|---|---|---|---|---|
| | **All-Cause Mortality** | **CVD Mortality** | **Cancer Mortality** | **Non-CVD/Non-cancer Mortality** |
| AMD status | | | | |
| No AMD | Reference | Reference | Reference | Reference |
| Any AMD | **2.14(1.39, 3.28)** | 1.42(0.68, 2.96) | 1.37(0.54, 3.51) | **2.40(1.43, 4.03)** |
| Early AMD | **2.05(1.33, 3.19)** | 1.27(0.51, 3.21) | 1.43(0.56, 3.65) | **2.40(1.37, 4.21)** |
| Late AMD | **2.99(1.59, 5.61)** | 1.78(0.92, 3.44) | 0.12(0.01, 1.07)[b] | **3.06(1.33, 7.05)** |

Abbreviations: AMD, age-related macular degeneration; CVD, cardiovascular disease; CI, confidence interval; HR, hazard ratio.

[a] Adjusted for age, gender, race, education, marital status, family income, smoking status, alcohol consumption, diabetes mellitus, hypertension, high cholesterol, BMI, high CRP, depressive symptom, coexisting ocular diseases, walking disability, self-rated health, and history of cardiovascular diseases.

[b] Adjusted for age and gender.

Boldface indicates statistical significance.
